# An outbreak report of the two autochthonous cases of airport malaria in Belgium in 2020

**DOI:** 10.1101/2021.09.29.21263834

**Authors:** Wim Van Bortel, Bea Van den Poel, Greet Hermans, Marleen Vanden Driessche, David Lerouge, Isra Deblauwe, Katrien De Wolf, Anna Schneider, Nick Van Hul, Ruth Müller, Leen Wilmaerts, Sophie Gombeer, Nathalie Smitz, Johanna Helena Kattenberg, Pieter Monsieurs, Anna Rosanas-Urgell, Marjan Van Esbroeck, Emmanuel Bottieau, Ula Maniewski-Kelner, Javiera Rebolledo

## Abstract

We report an outbreak investigation of two fatal cases of autochthonous *Plasmodium falciparum* that occurred in Belgium in September 2020. Various hypotheses of potential source of infection were investigated. Based on the collected information, the most likely route of transmission was through an infectious exotic *Anopheles* mosquito that arrived via the international airport of Brussels or the Military airport Melsbroek and infected the cases who lived at five kilometres from the airports. Based on a genomic analysis of the parasites collected from the two cases, the most likely origin of the *Plasmodium* was Gabon or Cameroon. Further, the parasites collected from the two Belgian patients were identical-by-descent, which supports the assumption that the two infections originated from the bite of the same mosquito, during an interrupted feeding. Despite these cases, airport malaria remains a rare event. Yet, it has significant implications, particularly for the patient, as delayed or missed diagnosis of the cause of illness often results in high rates of complications and mortality. Therefore, to prevent such severe or fatal outcomes, a number of public health actions are suggested including increased awareness among health practitioners especially those working in the vicinity of airports and increased surveillance of exotic mosquito species at airports.

## Background

Malaria is a protozoan disease caused by the *Plasmodium* parasite and transmitted by the bite of an infectious *Anopheles* mosquito. In Belgium, malaria is a travel-associated infection. Between 2016 and 2019, 327 to 420 cases were reported annually by the Belgian National Reference Laboratory. Due to the SARS-CoV-2 pandemic and the subsequent decrease of travel, the number of cases dropped to 179 in 2020. Cases due to local transmission are only sporadically reported in Belgium. In 1995 a cluster of six cases of airport malaria, including five cases of *Plasmodium falciparum* and one case of *Plasmodium ovale*, was reported from the international airport of Brussels, involving three airport employees and three visitors who did not travel abroad. One patient died [1]. In 1997 a possible case of port malaria was described in Ghent [2] and in 1998 a case of airport malaria was reported from the regional airport of Oostende [3]. In 2008 another case was notified in Brussels labelled as suitcase malaria [4]. In January 2015, a 74 year old women was diagnosed with *P. falciparum* malaria near Antwerp. The exact route of transmission has not been elucidated, but suitcase malaria was suspected [5].

The reporting of suspected autochthonous malaria is mandatory in Belgium. In case of suspected local transmission different hypotheses of potential source of infection need to be investigated [6]. First of all, travel related malaria, acquired in endemic zones, needs to be considered and excluded. Second, the investigation should assess the possibility of induced malaria, i.e. non-mosquito transmitted malaria for example through blood transfusion or nosocomial transmission. Third, introduced malaria, meaning the possibility of transmission by a local *Anopheles* species which infected itself by taking a blood meal on a gametocyte carrier (originating from endemic areas) needs to be examined. Finally, Odyssean malaria which results from the bite of an imported infectious exotic *Anopheles* mosquito can be considered.

Here we report the outbreak investigation of two fatal cases of autochthonous *P. falciparum* that occurred in the vicinity of the international airport Brussels and the military airport of Melsbroek in September 2020.

### Outbreak detection

#### Case 1

In the last week of September 2020 a women between 80-85 years old with a history of obesity and arterial hypertension presented to the emergency ward of the hospital with acute-onset diarrhoea and dyspnoea for one day. She had a documented fever (38.7 °C) at the time of admission. She was conscious and oriented. Laboratory investigation showed elevated inflammatory markers, metabolic acidosis, and liver and kidney failure. Blood cell count showed a thrombocytopenia of 88 × 10^9/L, but no anaemia. Despite antibiotic therapy and supportive treatment, the patient rapidly deteriorated and died of refractory shock in the night of 30 September. Blood cultures sampled at admission remained negative. Diagnosis of severe malaria was made retrospectively (parasitaemia at admission 8 %) when malaria was diagnosed in her husband (Case 2).

#### Case 2

Case two was an 80-85 years old man. He was admitted in the last week of September 2020 with a one-week history of cough, dyspnoea and fever. Upon hospital admission, the patient was conscious with a temperature of 38.2 °C. The patient was tachycardic (120/min), tachypneic (27/min) and required supplemental oxygen (9 L). Broad-spectrum antibiotic therapy was initiated. Laboratory investigation showed elevated C-reactive protein (up to 335 mg/L), neutrophilic leucocytosis with myeloid precursor cells and severe thrombopenia (24 × 10^9 /L) but no anaemia. Metabolic acidosis with lactate levels of 7.1 mmol/L and haemoglobinuria were present, as well as elevated D-dimers and lactate dehydrogenase. Because of haematological abnormalities, the blood sample was ‘flagged’ by the analyser (Sysmex XT4000i). Microscopic investigation by the laboratory technician revealed malaria parasites with a parasitaemia of 30 %. Immediately after diagnosis, intravenous quinine therapy was initiated. The patient rapidly developed respiratory failure, shock and acute kidney injury and was transferred within 24 h to the University Hospital. Upon transfer, therapy was switched to artesunate. Despite pharmacological hemodynamic support, continuous renal replacement therapy, correction of hypoglycaemia, metabolic acidosis further progressed and rhabdomyolysis developed. The patient died within 48 hours after transfer due to refractory shock and multiple organ failure.

Samples from both patients were sent to the malaria national reference laboratory at the Institute of Tropical Medicine, Antwerp (ITM) for confirmation. Parasite density quantification was assessed according to the WHO standards for microscopy [7]. Microscopy revealed *P. falciparum* trophozoites at a parasite density of 407.144 asexual parasites (AP)/µl (8.16 %) in Case 1 and 1 170 852 AP/µl (29.20 %) in Case 2. The presence of *P. falciparum* was confirmed with PCR using the method described in [8] with Ct values of 12.88 and 12.32 respectively.

Since travel history was absent for both cases, the cases were immediately reported to the regional health authorities as part of the mandatory notification when autochthonous malaria is suspected and triggered an outbreak investigation to assess the possible route(s) of transmission in order to put prevention and control measures in place if needed.

## Methods

### Overview of the outbreak investigation

The various hypotheses of potential source of infection were investigated. The travel history of the cases was carefully assessed (travel related malaria, hypothesis 1), as well as the transmission through blood-transfusion or nosocomial transmission (induced malaria, hypothesis 2). An entomological investigation was performed in and around the house of the cases in search for exotic and native *Anopheles* mosquitoes. Postal codes from patients with imported *P. falciparum* infections in the months of August and September 2020 were compared to the postal code of the couple in order to identify a possible index case for introduced malaria (hypothesis 3). Further, the hypothesis of Odyssean malaria (hypothesis 4) was assessed. In order to advance the investigation, whole genome sequencing (WGS) of the parasite genomes of the two samples collected from the cases was implemented to assess the most likely origin of the parasite.

### Entomological investigation

The entomological investigation searched for the presence of *Anopheles* mosquitoes in the house and garden of the cases, and in a wider area of 500 m around the house.

On 9 October, the house of the cases was inspected by two entomologists for the presence of adult mosquitoes using a mouth aspirator and a torch. The same day a search for larvae was conducted in the garden of the house and a BG Sentinel trap (Biogents) provided with BG Lure and CO_2_ was set-up. The trap content was collected one week later, on Friday 16 October. Additionally, a mosquito collected inside the house in the week after the death of the cases by one of the family members was also taken for species identification. In case the collected mosquitoes in the house belonged to the *Anopheles* genus, the head and thorax were tested for *Plasmodium* spp. using the PCR method described in [9].

On 12 October a mosquito larval collection was organized in a 500 m radius around the house of the cases. Different potential larval habitats were inspected including tree-holes, ponds, sewers, and any water holding containers that were present in the gardens of the neighbours and in the surroundings. In the nearby nature reserve ‘Torfbroek’ the marsh was inspected and in the forest ‘Hellebos’ tree holes and a pond were checked.

On 13 October, the outside area of the military airport of Melsbroek was inspected by two members of the Veterinary Service of Belgian Defence to check for larval habitats.

Adult mosquitoes and larvae were morphologically identified to genus level. Specimens of the *Anopheles* genus were further identified to species level [10, 11]. DNA barcoding was performed to confirm the morphological species identification [12].

### Genomic analysis of parasite origin

DNA extraction was performed on the blood samples of the two cases and WGS reads were generated on the Illumina NovaSeq platform. The analysis used the *P. falciparum* Community Project Pf6 [13] WGS database in addition to WGS data from Gabon [14] for comparison (for details see Supplemental Material). The geographical origin of the two *P. falciparum* isolates detected in Belgium was explored using a Principal Component Analysis (PCA), as implemented in the scikit-allele python module and using a Discriminant Analysis of Principal Components (DAPC) from the adegenet package in R [15]. The first analysis did not use a priori knowledge on the possible origin of the samples; the second calculated the discriminant components using country information (Supplemental Material).

The relatedness between the two Belgian isolates and samples nearest in the PCA was subjected to identity-by-descent (IBD) analysis, which indicates common ancestry between different strains, using the R-package isoRelate [16]. Nucleotide sequences are IBD if they are identical and inherited without recombination. isoRelate performs a pairwise relatedness-mapping on genomes from haploid isolates using a first order continuous time hidden Markov model allowing for multiple infections as often observed in *Plasmodium* infections. Pairwise comparisons between isolates from Belgium and countries in the identified cluster (Democratic Republic of Congo (DRC), Gabon, Cameroon, Nigeria, and Benin) were conducted using the same package and proportion of the genome sharing IBD was calculated.

## Results

### Epidemiological and clinical case investigation

None of the cases had a travel history outside Belgium since more than 50 years. The couple spent most of their time inside their home and only went to nearby shops. They did not get any blood or organ transfusion in the last five years and were not admitted to any hospital wards three years prior to the malaria episode, excluding transmission through substances of human origin and nosocomial transmission. In total 15 imported *P. falciparum* infections were reported in Belgium in August– September 2020. None of these cases lived close to the couple and could have been the source for infection of a local *Anopheles* mosquito. Cases were living at five kilometres from the international Airport Brussels and the military airport Melsbroek, therefore Odyssean malaria was suspected (Figure 1, Table 1).

**Figure 1.**
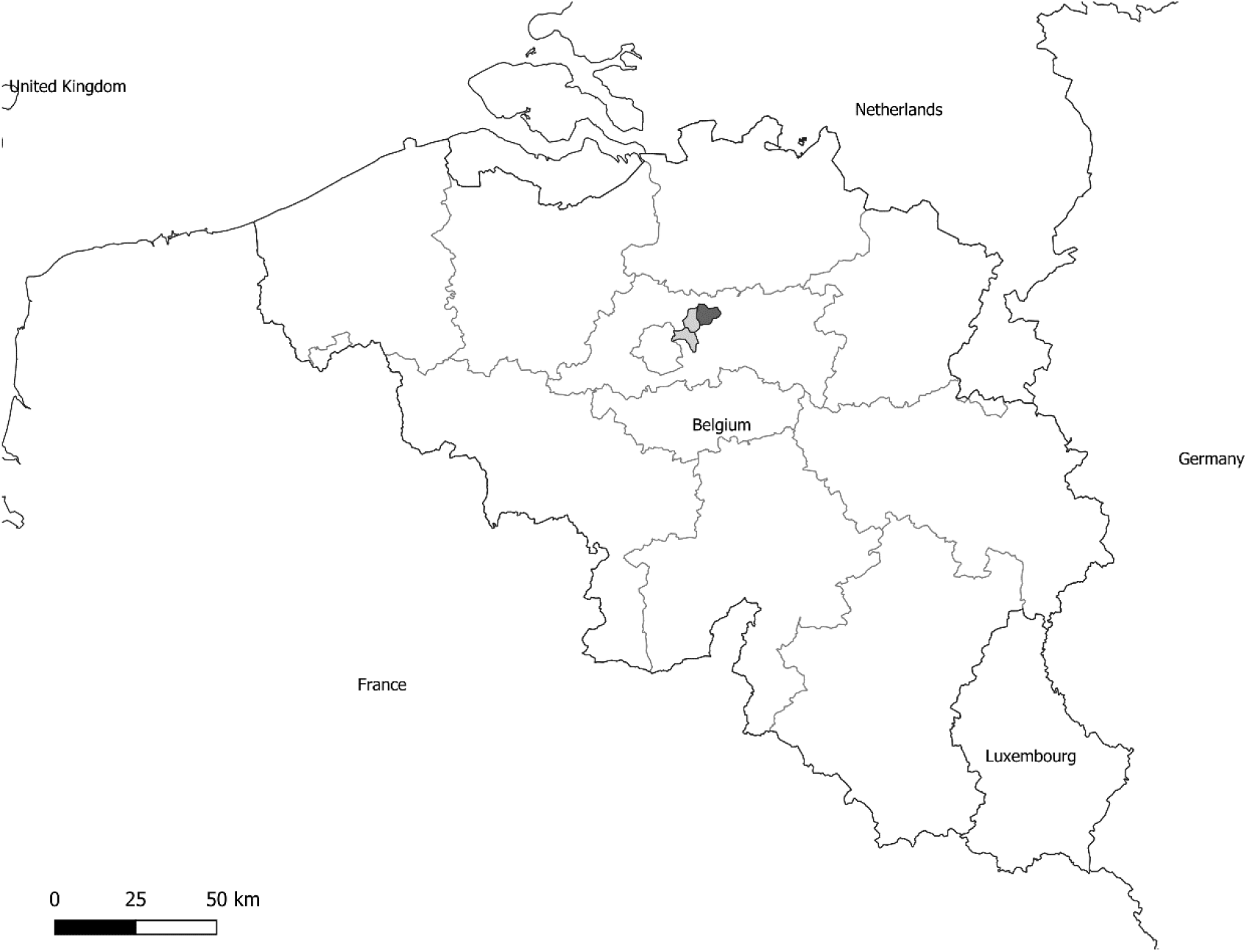
Location of the commune (dark grey) where the two autochthonous malaria cases were reported from in Belgium and the communes (light grey) where the international airport of Brussels and the military airport Melsbroek are located.

**Table 1.**
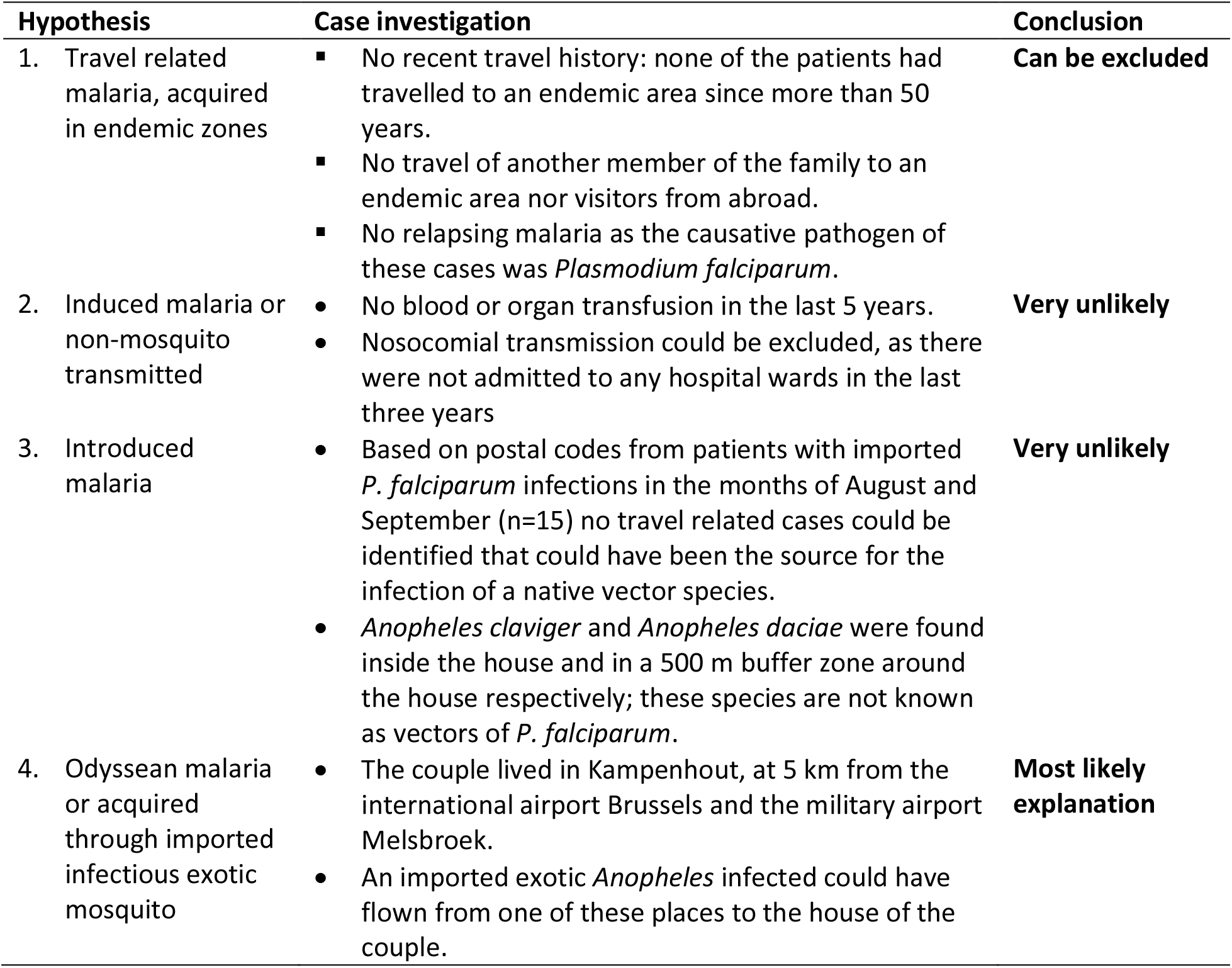
Overview of the outbreak investigations of two fatal autochthonous malaria cases in Belgium in 2020.

### Entomological investigation

In the house of the cases a single dead female mosquito was collected on the windowsill of the garage. The mosquito was identified as *Anopheles claviger* using morphology and DNA barcoding (GenBank accession number: MZ490542). The thorax and head portion of the female mosquito specimen was tested for the presence of the malaria parasite (*Plasmodium* spp.) which turned out to be negative. The mosquito collected by a family member was identified as *Culex pipiens / Cx. torrentium*. Using a mouth aspirator, one adult male of the native mosquito *Culex pipiens / Cx. torrentium* was collected from a rain barrel in the garden. The BG-Sentinel trap collected an additional ten specimens of *Culex pipiens / Cx. torrentium*. In the garden, a total of 37 containers of three different types of potential larval habitats were inspected, more specifically 30 plastic containers (such as flower pots, buckets and rain barrels), five plastic sheeting and two metal containers (e.g. drinking troughs). Most plastic containers and tarpaulins were positive for the presence of *Culex pipiens / Cx. torrentium*. No other mosquito species were found in the vicinity of the house.

In the 500 m buffer zone around the house of the cases, 23 gardens were visited and a total of 139 containers of 11 different types of potential larval habitats were inspected. In the gardens and the ‘Hellebos’ only *Culex pipiens / Cx. torrentium* larvae and pupae were collected. In the nature reserve ‘Torfbroek’ one pupa of the native mosquito *Anopheles daciae* was collected. Its identification was based on morphology and DNA barcoding (GenBank accession numbers: MZ490543 & MZ490544). Also the native *Culex pipiens / Cx. torrentium* and *Culiseta annulata* were collected in the marsh of this nature reserve.

On the site of the military airport Melsbroek, no potential larval habitats for mosquito were found and no mosquitoes were collected.

### Genomic analysis of parasite origin

In comparison to the database of *P. falciparum* genome sequences from across the world, the two Belgium isolates were placed in the large cluster with isolates from Africa and South America in the PCA, based on the genetic variation observed in those samples (Suppl. Figure 1A). In the PCA on a sub-set of the samples from Africa and South America, the Belgian isolates were placed near Western and Central African strains and closest to isolates from the interface between Western and Central Africa (Suppl. Figure 1B), but not the DRC (samples collected in Kinshasa). By enhancing the between-country variation using DAPC, the Belgian isolates were positioned closest to Gabon and Cameroon, and both isolates were predicted with the highest posterior probability (0.99) to belong to the cluster with samples from Gabon in the DAPC including samples from Western and Central Africa (Figure 2).

**Figure 2.**
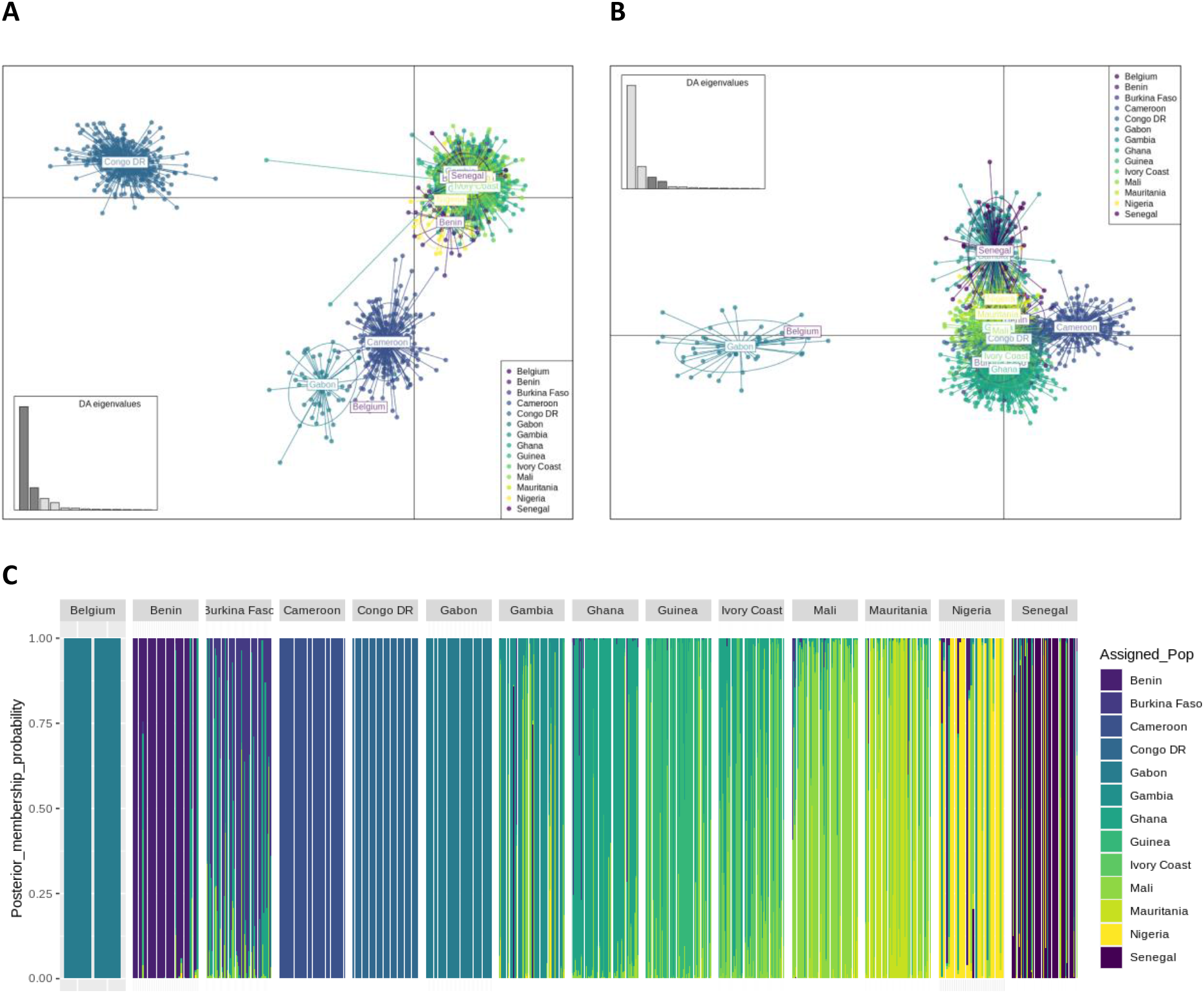
Discriminant analysis of principle components (DAPC). A) Scatter plot of DA eigenvalues 1 and 2, positioning Belgian isolates with Cameroon and Gabon; B) Scatter plot of DA eigenvalues 3 and 4, positioning Belgian isolates with Gabon; C) Composition plot of predicted posterior membership probability for Belgian isolates using DAPC with all Western and Central African isolates.

The parasite isolates collected in Belgium were 100 % IBD indicating that they were inherited from the same ancestor. However, little IBD was observed between the Belgian isolates and the selected samples from countries in Western and Central Africa, indicating no recent shared ancestry. The exception was one isolate from Gabon showing 2.9 % IBD with the Belgian isolates. This low IBD indicates a shared, although distant ancestry. However, in this part of Africa, there is high transmission and an intensely recombining parasite population, therefore it is expected that IBD proportions shared between isolates within the same country is not very high, and decreases over time [14]. The proportion of the genome in IBD in the pairwise comparisons within countries was 2.9 % [0.3-100 %] and larger than generally found in pairwise comparisons of isolates from different countries (1.8 % [0.68-8.0 %]).

### Flight information

Over the period 1 August– 15 September 2020, 471 flights – cargo and passengers flight combined – arrived from Africa at the international airport Brussels of which 102 direct flight originated from West Africa, 20 from Cameroon and none from Gabon. Compared to 2019, 65 % fewer flights arrived from Africa at the international airport Brussels in 2020. Despite this decrease, direct flights arrived from 14 African countries at the international airport Brussels in the period 1 August– 15 September 2019, whereas 20 African countries were connected via direct flight to the international airport Brussels in the same period in 2020 (Figure 3). Further most of the indirect flights had a stop-over in an African country (Figure 3). In the same period, 1 August– 15 September 2020, six direct and six indirect flights arrived from West Africa at the Military airport Melsbroek, but no flights arrived from Gabon nor from Cameroon.

**Figure 3.**
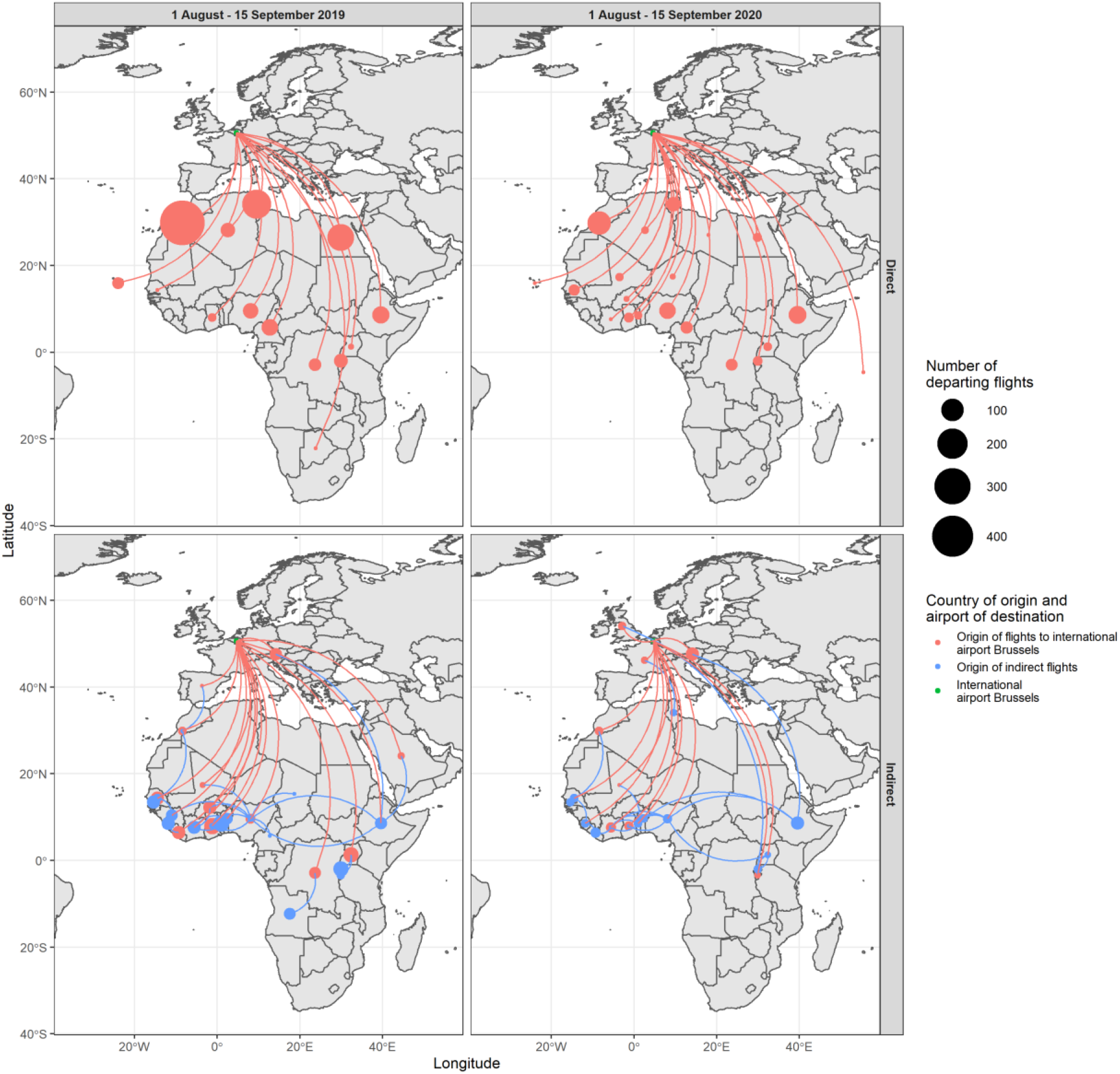
Overview of the number of direct and indirect flights from Africa to the international airport Brussels in the period 1 August–15 September 2019 and 2020.

### Outbreak control measures

As this transmission event was an isolated occurrence, the outbreak control measures included primarily communication to the population living in the surroundings of the cases. This included a press moment organised by the commune and information leaflets distributed to the population of the commune. Further, health professionals of the Region Flanders were informed about these cases to increase awareness. The outbreak investigation and the implementation of the control measures are the competences of the regional health authorities. At national level a risk assessment was made by the Risk Assessment Group coordinated by Sciensano and approved by the Risk Management Group. No further control measures were deemed necessary.

## Discussion

We described two fatal cases of autochthonous *P. falciparum* malaria in Belgium in 2020, which occurred at the beginning of the second Covid-19 wave in Belgium. Based on the information collected during the outbreak investigation the most likely route of transmission was through an infectious exotic *Anopheles* mosquito that arrived via the international airport of Brussels or the Military airport Melsbroek and infected the cases.

The entomological investigation in and around the house of the cases revealed the presence of two native *Anopheles* species, *An. claviger* and *An. daciae*. The former was a vector species of *Plasmodium vivax* in the eastern Mediterranean region [17]. The vector competence of *An. daciae* is not known as this species was only recently described [18] but the species could possibly be held responsible for *P. vivax* transmission that was attributed to *An. messeae* [17, 19]. Neither species is known as vector of *P. falciparum*. During the entomological investigation, we did not find *Anopheles plumbeus*, a competent *P. falciparum* vector [20] which was incriminated as vector in two autochthonous *P. falciparum* cases in Germany in 1997 [21]. Furthermore, the investigation of imported malaria cases in the vicinity of the cases did not reveal any travel associated malaria case which could have been identified as index case. Based on these observations it is very unlikely that local *Anopheles* mosquitoes can be held responsible for the local malaria cases.

One of the cases developed symptoms around 22 September. Given the incubation period of 8-12 days for *P. falciparum*, the transmission most likely took place in the second week of September. During that period Belgium experienced high temperatures [22], similarly as in 1995 when a cluster of six airport malaria cases were reported from the international airport of Brussels. Due to the Covid-19 pandemic, in 2020 compared to 2019 we observed a lower number of flights from the likely location of origin of the parasite which most likely resulted in a lower probability of entry of exotic mosquitoes through this import route. Yet, the hot spell could have favoured the survival of introduced exotic *Anopheles* mosquitoes. As the couple lived at five kilometres from the international airport Brussels and the military airport Melsbroek, an infectious exotic *Anopheles* mosquito could have survived and reached the house of the cases. In fact, the import of exotic *Anopheles* species does occur in Belgium. In 2017, a female *Anopheles pharoensis*, a malaria vector from Africa, was intercepted at the cargo airport of Liège during the Monitoring of Exotic Mosquito species Project [23, 24]. Also in the Netherlands exotic *Anopheles* species are sporadically intercepted at airports [23]. The isolates from the two Belgian patients were IBD, which supports the assumption that the two infections originated from the same exotic mosquito, during an interrupted feeding.

Odyssean malaria, also called airport, port or suitcase malaria, results from the bite of an infectious exotic mosquito imported into the EU in an aircraft or boat [25]. The conclusion of this possible route of infection is often based on excluding other hypothesis as shown in Table 1, since a direct observation of this event is very unlikely. Yet, the advanced genetic analysis provided additional elements to the investigation. The genetic signatures from the isolates from the cases indicated that they were most closely related to isolates from Gabon. Parasite subpopulation structure was observed in the dataset separating West, Central and South-Central African populations, and the Belgian strains clustered with the central region of Africa near Gabon and Cameroon. Predicting the country of origin from genomic data largely depends on the completeness of the parasite WGS database. In this context, major limitations existed in the available data in terms of (1) geographic representation – the more countries and sites within a country are represented in the genomic dataset, the higher the accuracy of predicting the origin; and (2) the time difference between the collection of the reference dataset and the collection of isolates for prediction, as parasite population structure changes over time due to frequent recombination. In the analyses the currently available data from online data sources were used. The majority of the samples were collected between 2007 and 2015 and only from few sites in a limited number of countries. The isolates from Gabon were collected in 2012, while the isolates from Cameroon were collected in 2013. Larger datasets of more recent isolates collected from multiple countries and areas within countries are required to enable a more precise prediction of the origin of imported isolates. Networks of collaborating research institutes and/or reference laboratories in endemic and non-endemic areas involved in isolate collection and/or malaria genomics and surveillance, as well as data sharing policies could contribute towards expanding the existing datasets.

In the period 1987-1995, 31 cases of airport malaria have been reported in Europe [26], including the cluster of six cases in Belgium in 1995 [1]. More recently, in 2019, two airport associated malaria cases were reported from Germany [27]. In 2020, besides the two cases in Belgium, also three airport malaria cases were reported from France [28]. Despite these occurrences, Odyssean malaria remains a rare event. Yet, it has significant implications, particularly for the patient, as delayed or missed diagnosis of the cause of illness often results in high rates of complications and mortality. Therefore, to prevent such severe or fatal outcomes, a number of public health actions could be considered. A good human surveillance is essential to understand the epidemiology of travel related malaria in Belgium and a timely notification to local health authorities whenever there is a suspicion of malaria is of paramount importance to rapidly put an outbreak investigation in place. Raising awareness among health care practitioners working near airports is essential as they should consider malaria as a differential diagnosis when laboratory and/or clinical features such as recurrent and unexplained fever are observed despite the absence of known potential exposure to (exotic) mosquitoes or travel history. Further, the observation also points to the need for vigilance at airports for import of exotic mosquito species, the necessity for continuous monitoring in and around airports as well as the need for appropriate control of mosquitoes at airports, including the luggage and cargo spaces, and in airplanes [29], and this particularly during summer season.

## Data Availability

This is an outbreak investigation and all data are presented and available in the manuscript

## Ethics declaration

This case report was notified to the Chairperson of the Institutional Review Board of the ITM on 17/06/2021. It was exempted from formal ethics approval, provided that consent was provided by the family members.

## Acknowledgements

We would like to thank the family members of the cases who facilitated the entomological investigations in the house and garden and who consented to this publication, as well as the people in the neighbourhood that provided access to their gardens during the investigations.

## Funding statement

The Institute of Tropical Medicine implements exotic mosquito monitoring in Belgium through the projects The MEMO and MEMO+ project (2017 – 2020) which is funded by the Flemish, Walloon and Brussels regional governments and the Federal Public Service (FPS) Public Health, Food Chain Safety and Environment in the context of the National Environment and Health Action Plan (NEHAP) (Belgium). The entomological outbreak investigation was also financially supported by the “Agentschap Zorg en Gezondheid”. The Outbreak Research Team of the Institute of Tropical Medicine is financially supported by the Department of Economy, Science and Innovation of the Flemish government. The Barcoding Facility for Organisms and Tissues of Policy Concern (BopCo - http://bopco.myspecies.info/) is financed by the Belgian Science Policy Office (Belspo) as Belgian federal in-kind contribution to the European Research Infrastructure Consortium “LifeWatch”. The national reference laboratory of the Institute of Tropical Medicine is partially supported by The Federal Public Service Health, Food Chain Safety and Environment through a fund within the Health Insurance System. The genomic investigation was supported by Malariology Unit Funds to Rosanas-Urgell. The funders were not involved in the analysis nor the interpretation of the data.

## Author contribution

Wim Van Bortel initiated the entomological outbreak investigation, compiled the different aspects of the investigation and drafted the manuscript. Bea Van den Poel, Greet Hermans, Marleen Vanden Driessche, and David Lerouge did the initial clinical and laboratory investigation and Marjan Van Esbroeck provided the laboratory confirmation as head of the Clinical Reference laboratory of Belgium. Emmanuel Bottieau and Ula Maniewski-Kelner provided clinical expertise during the outbreak investigation. Isra Deblauwe, Katrien De Wolf, Anna Schneider, Nick Van Hul, Ruth Müller and Leen Wilmaerts implemented the entomological outbreak investigation. Sophie Gombeer and Nathalie Smitz were responsible for the molecular identification of the collected mosquitoes. Johanna Helena Kattenberg, Pieter Monsieurs and Anna Rosanas-Urgell performed the genomic analysis of parasite origin. Javiera Rebolledo verified the malaria surveillance data to compare postal codes and was responsible for the risk assessment at federal level. All authors contributed to the revision of the draft manuscript and approved the manuscript.

## Notes

### Competing Interest Statement

The authors have declared no competing interest.

### Author Declarations

This case report was notified to the Chairperson of the Institutional Review Board of the ITM on 17/06/2021. It was exempted from formal ethics approval, provided that consent was provided by the family members. We would like to thank the family members of the cases who facilitated the entomological investigations in the house and garden and who consented to this publication

