## Supplementary Material 1 for "An outbreak report of the two autochthonous cases of airport malaria in Belgium in 2020"

**The geographical origin of the two *Plasmodium falciparum* isolates detected.**

***DNA extraction.***

≥10ml of venous blood in EDTA was obtained from each case and centrifuged at 1500 rpm for 5 min. Plasma was removed and the red blood cell pellet resuspended in PBS at 10% hematocrit (Htc). White blood cells were removed using cellulose columns as described previously [1]. DNA was extracted with QIAmp DNA mini blood kit (Qiagen) (using 200 µl 50 % Htc per case) following the manufacturer's instructions. Extracted DNA was eluted in 50 µL of elution buffer and 856 ng and 111 ng of DNA of the male and female cases respectively, was sent to GenomeScan (Leiden, the Netherlands) for sequencing.

***Analysis of Belgian samples***

Paired sequencing reads of 2x150 bp were generated on the Illumina NovaSeq platform. Reads were aligned to the 3D7 *P. falciparum* reference genome as available on the PlasmoDB website (version 48) [2, 3] using BWA (version 0.7.17) with seed length set to 50. Quality control was performed using FastQC [4] and FastQ Screen [5] and showed only a minor contamination with human DNA (lower than 3 % of the reads for both samples). Only properly paired reads with a mapping quality higher than 30 were selected using SAMtools [6]. Duplicate reads were removed using the RemoveDuplicates command in the Picard software [7]. SNP calling was performed using the GAKT pipeline [8]. Only those positions were retained for which a genetic variant is described in the *Plasmodium falciparum* database obtained from MalariaGEN.net.

***Database of Plasmodium falciparum***

A catalogue (version 6.0) of genetic variants was downloaded from the *Plasmodium falciparum* Community Project (MalariaGEN.net), which contains genotype calls of 7113 strains, each of them genotyped using the GATK pipeline resulting in 1,045,910 polymorphic coding SNPs across the genome. For the samples originating from Gabon, which are not included in this catalogue, sequencing data were downloaded from the NCBI Sequence Read Archive based on the accession numbers provided in [9], and consecutively subjected to the same pipeline as described above for the Belgian samples. All genetic variants were combined using the merge command of the BCFtools. Biallelic SNP variants in the core genome region only [10] were filtered and subjected to LD pruning using the 'locate_unlinked' function from the python package scikit-allel [11]. LD-pruning was performed in several iterations until no more variants were removed.

***Data analysis***

The PCA was applied to two data sets: (1) *Plasmodium* samples from all countries which passed Quality Control; and (2) isolates from West and Central Africa, and South America. While PCA clusters the different samples without foreknowledge, Discriminant Analysis of Principal Components (DAPC) [12] calculates the discriminant components using the country information in such a way that samples originating from the same country will group together, while it tries simultaneously to maximize the distance with samples from other countries. DAPC was performed using the R library Adegenet on the latter data set, excluding the South American countries and Belgian isolates using 200 principal components (PCs) and 12 discriminants (determined through cross-validation). Based on those DAPC results, posterior probabilities for the Belgian samples were calculated, which gives for every country the probability whether either of the Belgian samples has a similar genetic profile as the isolates from those countries. For visualization DAPC was also conducted including the Belgian isolates with 200 PCs and 13 discriminants in the analysis.

**Supplementary material. Figure 1.Principle component analysis (PCA)** of biallelic variants in the core genome region of isolates from A) across the whole world, and B) Western, Central and South-Central African countries. The first two components are depicted on the left, components 3 and 4 on the right of the figure. Isolates collected in Belgium are indicated as red dots, and cluster with isolates from Central Africa. WAF = West Africa, EAF = East Africa, WSEA = West South-East Asia, ESEA = East South-East Asia, OCE = Oceania, SAM = South America, SAS = South Asia, CAF = Central Africa.

**A.**

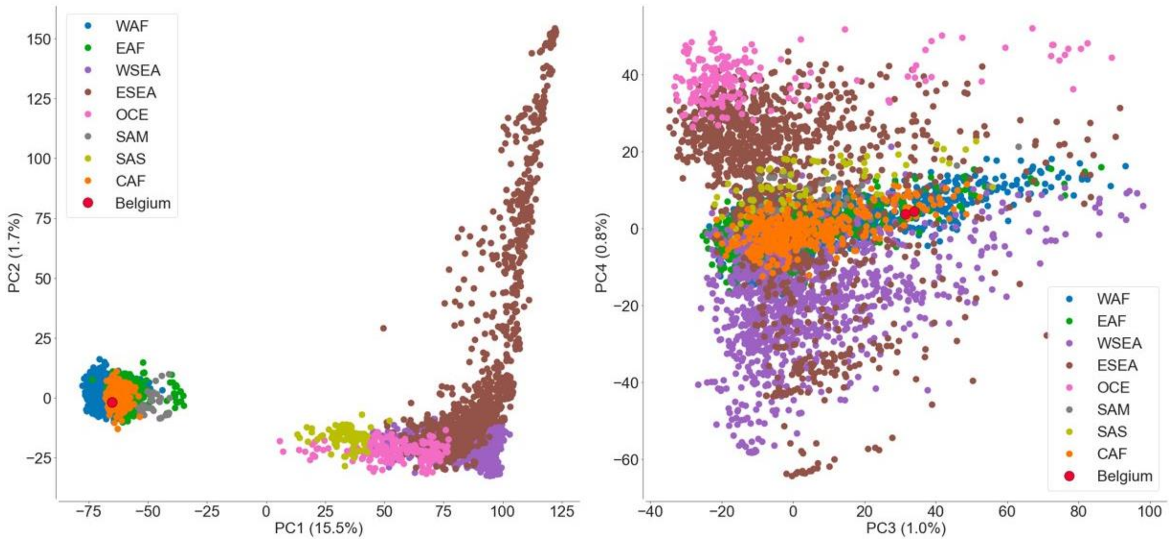


**B.**

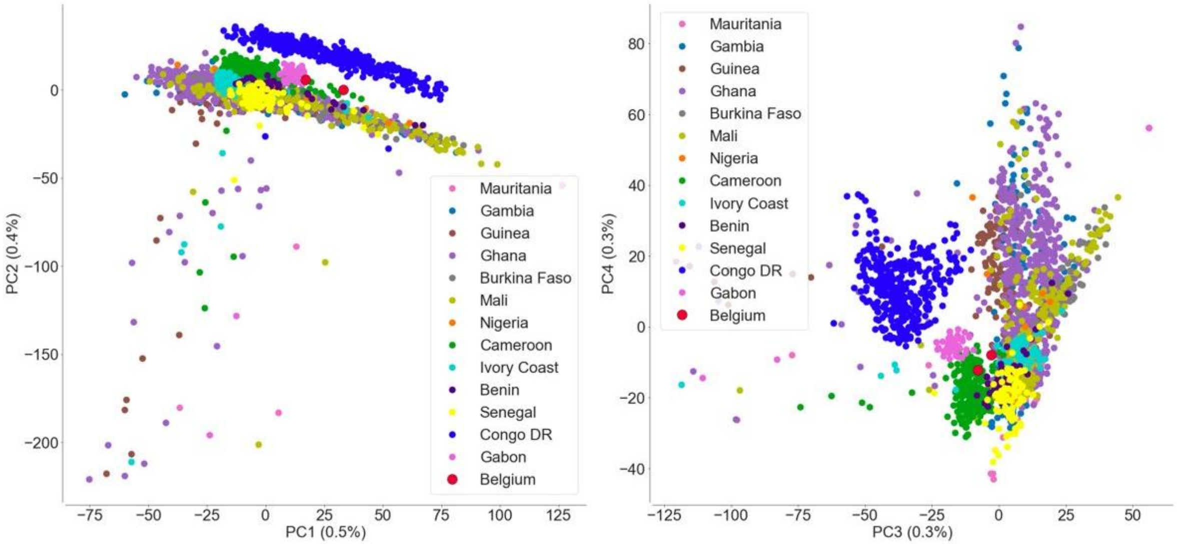

**References.**
